# Purchase, consumption, and ownership of chickens and chicken products among households in Maputo, Mozambique: A cross-sectional study

**DOI:** 10.1101/2024.05.14.24307337

**Authors:** Kayoko Shioda, Frederica Lamar, Hermógenes Neves Mucache, Anushka Reddy Marri, Jhanel Chew, Karen Levy, Matthew Freeman

**Affiliations:** Department of Global Health, School of Public Health, Boston University, Boston, MA; Center on Emerging Infectious Diseases, Boston University, Boston, MA; Gangarosa Department of Environmental Health, Emory University Rollins School of Public Health, Atlanta, GA; Veterinary Faculty, Universidade Eduardo Mondlane, Maputo, Mozambique; Department of Epidemiology, School of Public Health, Boston University, Boston, MA; Department of Environmental and Occupational Health Sciences, University of Washington School of Public Health, Seattle, WA

**Keywords:** chickens, foodborne disease, food safety, Maputo, Mozambique, population-based survey, poultry production

## Abstract

**Background:** Chickens are an important source of animal protein, nutrition, and income in many low- and middle-income countries (LMICs). They are also a major reservoir of enteropathogens that contribute to the burden of illnesses among children. Food systems present a risk for transmission of enteropathogens from poultry to humans, but there is a lack of population-level data on the pattern of purchase, ownership, and consumption of live chickens and their products in LMICs to better characterize that risk.

**Methods:** To assess chicken purchase, ownership, and consumption practices, we conducted a population-based survey using a structured questionnaire in Maputo, Mozambique in 2021. Multi-stage cluster sampling was used to obtain a representative sample of households in our study area. To minimize sampling bias and ensure a representative sample, we applied survey weighting using district-level population data and estimated weighted population-level values.

**Results:** Heads of 570 households in Maputo completed our survey. Approximately half of these households purchased broiler chicken meat (weighted percentage of households: 44.8%) and eggs (46.5%) in the previous week of the survey date, while indigenous chicken meat was less popular (1,950, 1.1%). The most common source of chicken products was corner stores (i.e., small convenience shops on streets), followed by wet markets. Live chickens were raised by 15.6% of households, and chicken feces were observed on the floor or ground at the majority of these households during house visits.

**Discussion:** Our findings suggest that poultry provides a major source of animal protein in this setting. With the predicted growth of poultry farming in LMICs in the coming decades, ensuring food safety at the primary sources of chicken products (corner stores and wet markets) in urban areas will be critical to mitigate health risks.

## INTRODUCTION

Chickens are a primary reservoir for high-burden zoonotic enteropathogens, in particular *Campylobacter jejuni/coli*, *Salmonella* spp., and Cryptosporidium.^1,2^ The 2010 Global Burden of Disease Study estimated that these three pathogens together were associated with more than 20 million disability-adjusted life years (DALYs) across all ages globally.^3^ Early colonization with these pathogens, specifically *Cryptosporidium*, are associated with growth shortfalls.^4–6^

Zoonotic enteropathogens can be transmitted to humans via multiple routes. Children and infants may get infected by ingesting chicken feces directly or via contaminated soils and surfaces.^7–9^ Evidence suggests that children who had household exposure to poultry are at an increased risk of diarrhea^10,11^ and anemia.^12^ Food systems, which are often largely unregulated in low- and middle-income countries (LMICs),^13^ also pose a major risk of transmission of zoonotic enteropathogens from poultry to humans. In high-mortality countries in the African region, 57% of *Campylobacter* spp. infections are attributed to foodborne transmission.^14^ In our previous work, *Campylobacter jejuni/coli* and *Salmonella* spp. were detected in chicken carcasses and chicken fecal samples collected along the poultry production system, such as small-scale farms and wet markets in Maputo, Mozambique.^15^ Indigenous chickens raised in backyards in Maputo also carried *C. jejuni/coli* and *Salmonella* spp.^15^

Chickens are promoted as a development strategy in LMICs and serve as a primary source of animal protein and income in many areas.^16^ Poultry is the fastest growing livestock subsector, and poultry meat and egg production have grown by 250% over the past 30 years in LMICs.^1^ Small-scale poultry production is increasing in resource-limited areas as a means of providing nutrition, income, and food security for households,^17^ contributing to multiple Sustainable Development Goals (SDGs), such as no poverty, zero hunger, and decent work and economic growth.^18^ Poultry farming also contributes to the empowerment of the youth and women.^19^ In Mozambique, the agricultural sector contributes 24% to its gross domestic product,^20^ and chickens make up nearly half of small and medium farms for livestock production.^21^ The consumption of poultry meat is expected to increase from 34.2 thousand metric tons in 2000 to 127.8 in 2030 in Mozambique,^1^ highlighting the importance of addressing food safety issue of poultry products.

Improved characterization of the pattern of purchase, ownership, and consumption of live poultry and their products would help quantify the magnitude of potential poultry-associated health risks to support efforts to promote food safety in LMICs. While benefits and risks of animal-sourced proteins have been assessed,^22,23^ to our knowledge, there have been few population-based studies on poultry use and consumption in LMICs^13^ and none in Mozambique.^24–26^ Such type of population-level data could help to further evaluate the competing risks and benefits of poultry production and identify effective control measures to reduce enteropathogen infection in LMICs. Such data would support better quantitative models to understand risks of various foodborne pathways of infection. To address this gap, we conducted a population-based survey in Maputo, Mozambique in 2021.

## MATERIALS AND METHODS

### Study background

We conducted a cross-sectional, population-based survey to estimate chicken and egg purchase and consumption within Maputo, the capital city of Mozambique, as well as to describe practices related to chicken rearing (e.g., litter disposal) and food handling. In Mozambique, chicken consumption and production have grown from approximately 56,000 tons of production and 61,000 tons of consumption in 2013 to 89,000 tons of production and 91,000 tons of consumption in 2017.^27^ Nine in-country hatcheries provide 46.5 million day-old chicks annually.^28^ Maputo, with a population of approximately 1.1 million,^29^ has a growing poultry sector.^28,30^

### Study objectives and survey design

We recruited households across five districts of Maputo and employed a structured questionnaire among heads of household between May and June 2021. A household was defined as a person or group of related or unrelated persons who usually live together in the same dwelling unit(s), who have common cooking and eating arrangements, and who acknowledge one adult member as head of household.

The key indicators of interest were the ownership of live chickens in the household, the frequency and source of purchase of live chickens and their products, and the frequency of consumption of poultry products. Information was collected for each type of poultry products (meat and egg) as well as the following type of chickens: broiler chickens (i.e., chickens specifically bred to provide meat), layer chickens (i.e., hens specifically bred to provide eggs), and indigenous chickens (i.e., local free-ranging chickens). We asked about locations where households purchased chicken products, such as wet markets,^31^ corner stores (i.e., small convenience shops on streets), supermarkets, directly from farmers, or from families, friends, and neighbors. For households with children under five years of age, we asked about the corresponding information for children. We asked about households’ activities that may affect the risk of exposure to chicken feces, such as applying chicken litter to vegetable gardens, keeping chickens inside of the home, and whether their young household members help with these activities. We also conducted structured spot check observations of household characteristics (e.g., feces on the floor). More details of the questionnaire and observational component of the survey can be found in the Supplementary Methods.

### Districts and neighborhoods

Data were collected in five of seven Maputo City districts (Nlhamankulu, KaMaxakeni, KaMavota, KaMubukwana, KaMpfumu, and KaTembe). We did not collect data in the remaining two districts (KaNyaka or KaMpfumo) because KaNyaka is an island representing 0.5% of the population, having no broiler farms and very low production of indigenous chickens. KaMpfumu is the city-center, having multi-story residential buildings, and does not allow the production or raising of chickens.

### Sampling households

We used multi-stage cluster sampling to obtain a representative sample of our study area to estimate the proportion of households that consumed poultry in the previous week. Assuming a conservative estimate of 50%, a confidence level of 95%, an alpha of .05, a relatively low intra-class correlation coefficient of .02 (no data on this parameter are available), and a logistically relevant number of households per cluster that our team could reach in one day (20), we estimate a minimal sample size of 531 in 27 clusters. As such, we ended up with a sample size of 540 households (=27 clusters x 20 households) (Supplementary Table 1). Using a random number generator, we selected neighborhoods from a list of all neighborhoods within each district (Supplementary Table 1). We then sampled two clusters within each of the sampled neighborhoods. Geographic boundaries for neighborhoods were delineated in Google Earth Pro. Two, randomly selected 300m x 300m grids were overlaid within the boundaries of each neighborhood using R (R Center for Statistical Computing; Vienna, Austria). Each of the four enumerators was randomly assigned to a corner of the grid and walked toward a direction (North, South, East, West) selected by a random number generator app, and recruited households in the selected direction. Starting at the designated point, enumerators selected every tenth household for participation. If compounds, having multiple homes within one lot, were selected, each individual household was counted. Prior to the start of data collection, enumerators piloted the survey in two neighborhoods that were not included in the survey sample.

### Sample weights

Survey weighting was conducted using district-level population data to minimize sampling bias and to provide a representative sample. The probability of selection in each district was calculated by dividing the number of households that completed the survey in each district by the estimated number of total households in the corresponding district. As data on the number of households in each district was not available, we calculated the estimated number of total households in each district by dividing the population size of each district by the average household size. The design weight for each participating household was the inverse of their probability of selection. These survey weights were then used to calculate the weighted total population counts, average, and standard error with the ‘survey’ package in R.^32^

### Ethics

The Institutional Review Board at Emory University (IRB00108546) and the Research Council to the Veterinary Faculty at Eduardo Mondlane University determined that this research was exempt from human subjects review, and the Municipality of Maputo (Reference number 754/SG/426/GP/2019) authorized this research. Prior to data collection, the study’s purpose and participant rights were explained in Portuguese, and participants provided verbal informed consent.

## RESULTS

### Characteristics of surveyed households

A total of 570 households completed the survey. The probability of selection was 0.31% in KaMavota, 0.31% in KaMaxaquene, 0.32% in KaMubukwana, 1.11% in KaTembe, and 0.29% Nlhamakulu. Approximately half of the households that completed the survey had at least one child under five years of age (weighted percentage: 44.2%) (Table 1). The weighted average household size was 5.7 (standard error [SE] 0.1), and the weighted average number of children under five was 1.4 per household (SE 1.0).

**Table 1.** Characteristics of households that completed the survey in May-June 2021, Maputo, Mozambique.

|  | Unweighted |  | Weighted |  |
| --- | --- | --- | --- | --- |
|  | mean (SD) |  | mean (SD)<sup>1</sup> |  |
| Characteristic* | All households<br>(N =570) | Households<br>with Children<sup>2</sup><br>(N =252) | All households<br>(N =175143) | Households<br>with Children<br>(N = 77372) |
| Size of household, mean<br>(SD) | 5.7 (2.8) | 6.8 (2.9) | 5.7 (2.9) | 6.8 (3.0) |
| Number of children in<br>households, mean (SD) | -- | 1.4 (0.9) | -- | 1.4 (1.0) |

<sup>1</sup>Survey weighting was conducted using district level population data. The design weight for each participating household was the inverse of their probability of selection in the corresponding district.
<sup>2</sup>Children under 5 years of age.
Abbreviations: SD, standard deviation.

### Purchase and consumption of chicken products (weighted analysis)

Approximately half of the households purchased broiler chicken meat (44.8%) and eggs (46.5%) in the previous week of the survey date, while indigenous chicken meat was less commonly purchased (1.1%) (Table 2). Similar numbers were reported among households with children under five. Households reported relatively high consumption of broiler meat (62.4%) and eggs (59.0%) in the previous week of the survey date. Similar to the purchase pattern, consumption of indigenous chicken meat was not common (3.1%). The same pattern was observed among households with children under five. Among children under five in households that consumed chicken products, the weighted average weekly frequency of consumption was 2.4 days for broiler chicken meat, 1.8 days for indigenous chicken meat, and 2.7 days for eggs.

**Table 2.**
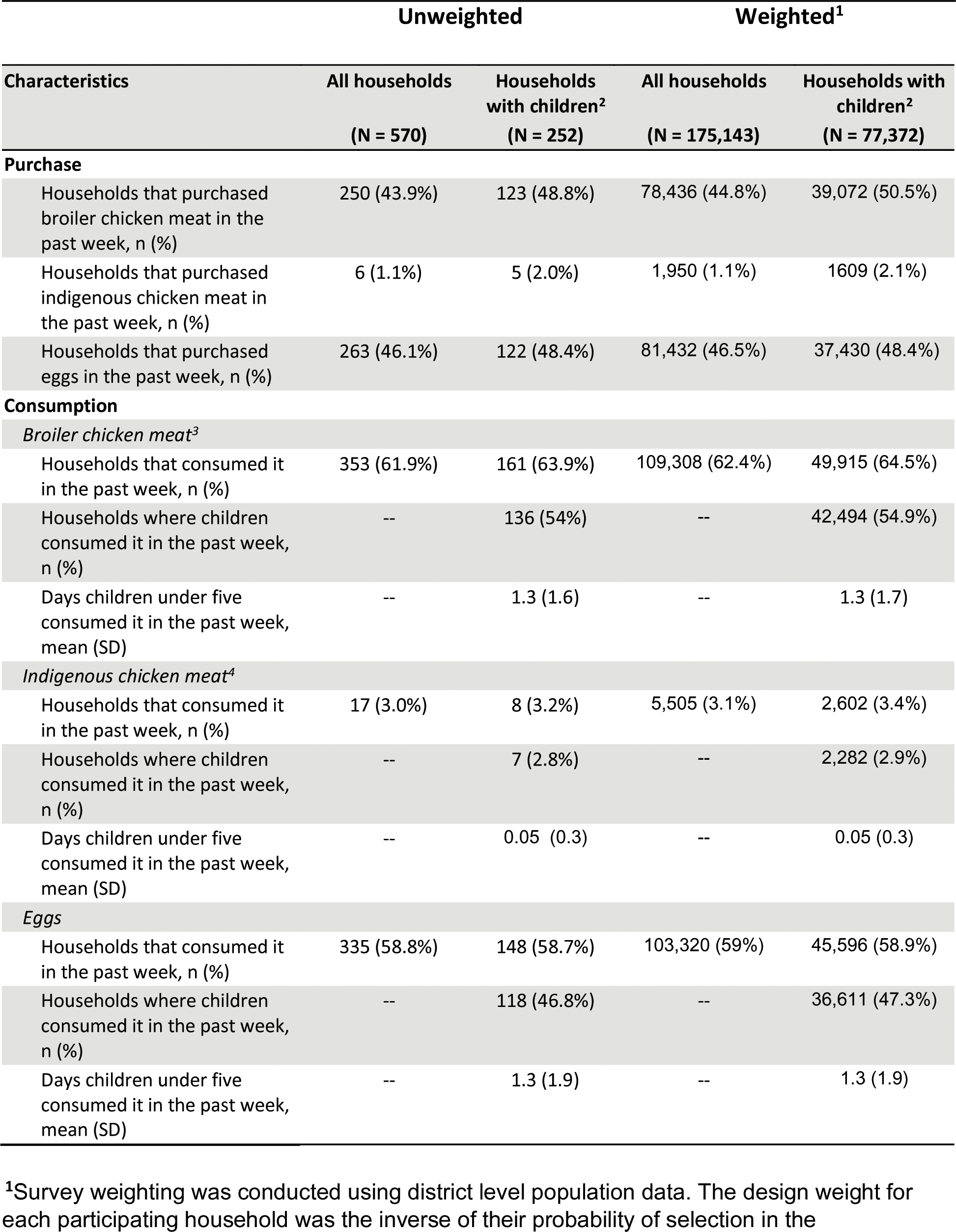
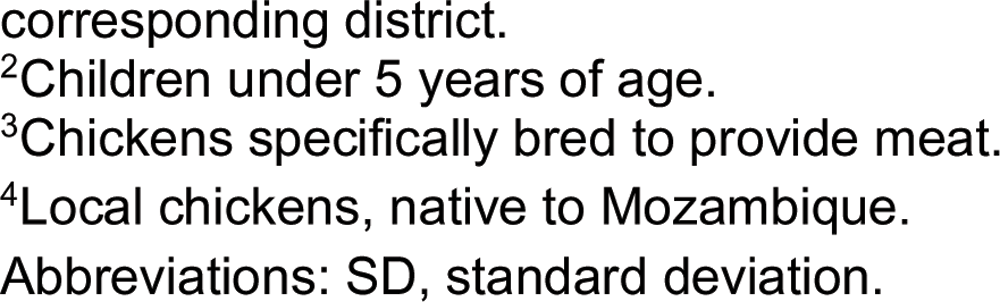
Purchase and consumption of poultry products in the past week of the survey date in May-June 2021 in Maputo, Mozambique.

| Characteristics | Unweighted |  | Weighted<sup>1</sup> |  |
| --- | --- | --- | --- | --- |
|  | All households<br>(N = 570) | Households with children<sup>2</sup><br>(N = 252) | All households<br>(N = 175,143) | Households with children<sup>2</sup><br>(N = 77,372) |
| **Purchase** |  |  |  |  |
| Households that purchased broiler chicken meat in the past week, n (%) | 250 (43.9%) | 123 (48.8%) | 78,436 (44.8%) | 39,072 (50.5%) |
| Households that purchased indigenous chicken meat in the past week, n (%) | 6 (1.1%) | 5 (2.0%) | 1,950 (1.1%) | 1609 (2.1%) |
| Households that purchased eggs in the past week, n (%) | 263 (46.1%) | 122 (48.4%) | 81,432 (46.5%) | 37,430 (48.4%) |
| **Consumption** |  |  |  |  |
| *Broiler chicken meat<sup>3</sup>* |  |  |  |  |
| Households that consumed it in the past week, n (%) | 353 (61.9%) | 161 (63.9%) | 109,308 (62.4%) | 49,915 (64.5%) |
| Households where children consumed it in the past week, n (%) | -- | 136 (54%) | -- | 42,494 (54.9%) |
| Days children under five consumed it in the past week, mean (SD) | -- | 1.3 (1.6) | -- | 1.3 (1.7) |
| *Indigenous chicken meat<sup>4</sup>* |  |  |  |  |
| Households that consumed it in the past week, n (%) | 17 (3.0%) | 8 (3.2%) | 5,505 (3.1%) | 2,602 (3.4%) |
| Households where children consumed it in the past week, n (%) | -- | 7 (2.8%) | -- | 2,282 (2.9%) |
| Days children under five consumed it in the past week, mean (SD) | -- | 0.05 (0.3) | -- | 0.05 (0.3) |
| *Eggs* |  |  |  |  |
| Households that consumed it in the past week, n (%) | 335 (58.8%) | 148 (58.7%) | 103,320 (59%) | 45,596 (58.9%) |
| Households where children consumed it in the past week, n (%) | -- | 118 (46.8%) | -- | 36,611 (47.3%) |
| Days children under five consumed it in the past week, mean (SD) | -- | 1.3 (1.9) | -- | 1.3 (1.9) |

<sup>1</sup>Survey weighting was conducted using district level population data. The design weight for each participating household was the inverse of their probability of selection in the
corresponding district.
<sup>2</sup>Children under 5 years of age.
<sup>3</sup>Chickens specifically bred to provide meat.
<sup>4</sup>Local chickens, native to Mozambique.
Abbreviations: SD, standard deviation.

### Source of chicken products and live chickens (weighted analysis)

The most common location where households purchased broiler chicken meat in the previous week of the survey date was corner stores (56.7% of households that purchased broiler chicken meat), followed by wet markets (17.5%), and directly from farmers (16.8%) (Table 3. Supplementary Table 2-4). Similarly, the primary source of eggs was corner stores (61.5% of households that purchased eggs), followed by markets (16.0%), and directly from farmers (8.4%). Live broiler chickens were most often purchased directly from farmers (63.4% of households that purchased live broiler chickens), from markets (19.4%), or from family, friends, or neighbors (10.8%). Live indigenous chickens were most commonly purchased from family, friends, or neighbors (51.2% of households that purchased live indigenous chickens).

**Table 3.**
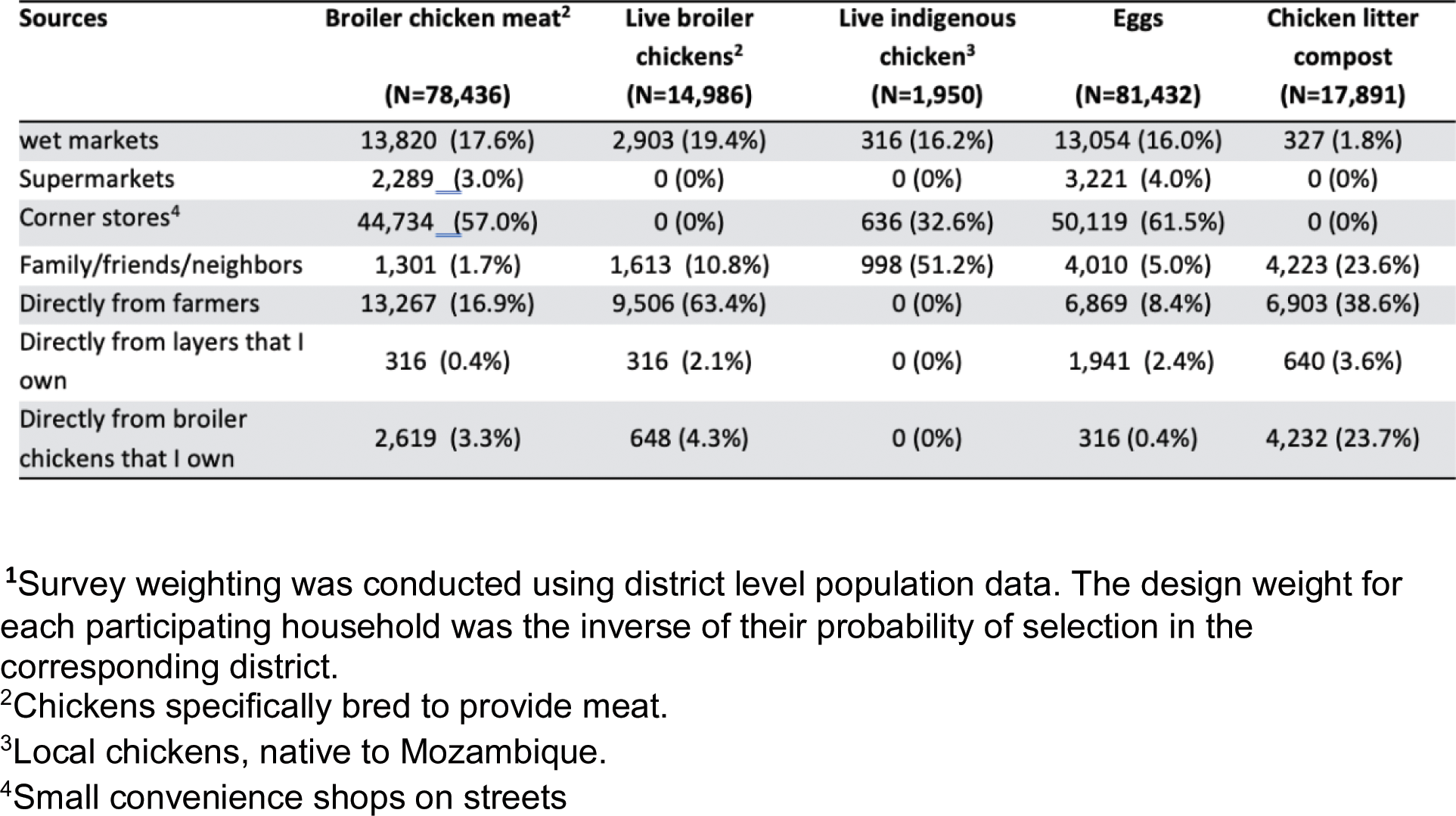
Source of live chickens, chicken meat, eggs, and chicken litter compost among households that purchased these products in the past week of the survey date in Maputo, Mozambique in May-June 2021 (weighted statistics^1^).

| Sources | Broiler chicken meat<sup>2</sup><br>(N=78,436) | Live broiler chickens<sup>2</sup><br>(N=14,986) | Live indigenous chicken<sup>3</sup><br>(N=1,950) | Eggs<br>(N=81,432) | Chicken litter compost<br>(N=17,891) |
| --- | --- | --- | --- | --- | --- |
| wet markets | 13,820 (17.6%) | 2,903 (19.4%) | 316 (16.2%) | 13,054 (16.0%) | 327 (1.8%) |
| Supermarkets | 2,289 (3.0%) | 0 (0%) | 0 (0%) | 3,221 (4.0%) | 0 (0%) |
| Corner stores<sup>4</sup> | 44,734 (57.0%) | 0 (0%) | 636 (32.6%) | 50,119 (61.5%) | 0 (0%) |
| Family/friends/neighbors | 1,301 (1.7%) | 1,613 (10.8%) | 998 (51.2%) | 4,010 (5.0%) | 4,223 (23.6%) |
| Directly from farmers | 13,267 (16.9%) | 9,506 (63.4%) | 0 (0%) | 6,869 (8.4%) | 6,903 (38.6%) |
| Directly from layers that I own | 316 (0.4%) | 316 (2.1%) | 0 (0%) | 1,941 (2.4%) | 640 (3.6%) |
| Directly from broiler chickens that I own | 2,619 (3.3%) | 648 (4.3%) | 0 (0%) | 316 (0.4%) | 4,232 (23.7%) |

<sup>1</sup>Survey weighting was conducted using district level population data. The design weight for each participating household was the inverse of their probability of selection in the corresponding district.
<sup>2</sup>Chickens specifically bred to provide meat.
<sup>3</sup>Local chickens, native to Mozambique.
<sup>4</sup>Small convenience shops on streets

### Ownership and household management of live chickens (weighted analysis)

Live chickens were raised by 15.6% of households at the time of survey (Table 4). The ownership of live chickens was less common among households with children under five (13.8%). The weighted average (range) number of chickens owned by a household at the time of survey was 7 (1-260) for broiler chickens, 5 (1-35) for indigenous chickens, and 10 (2-146) for layers (Table 4). It was more common for households to raise live chickens solely for personal consumption (13.7%) than for sale (1.3%).

**Table 4.**
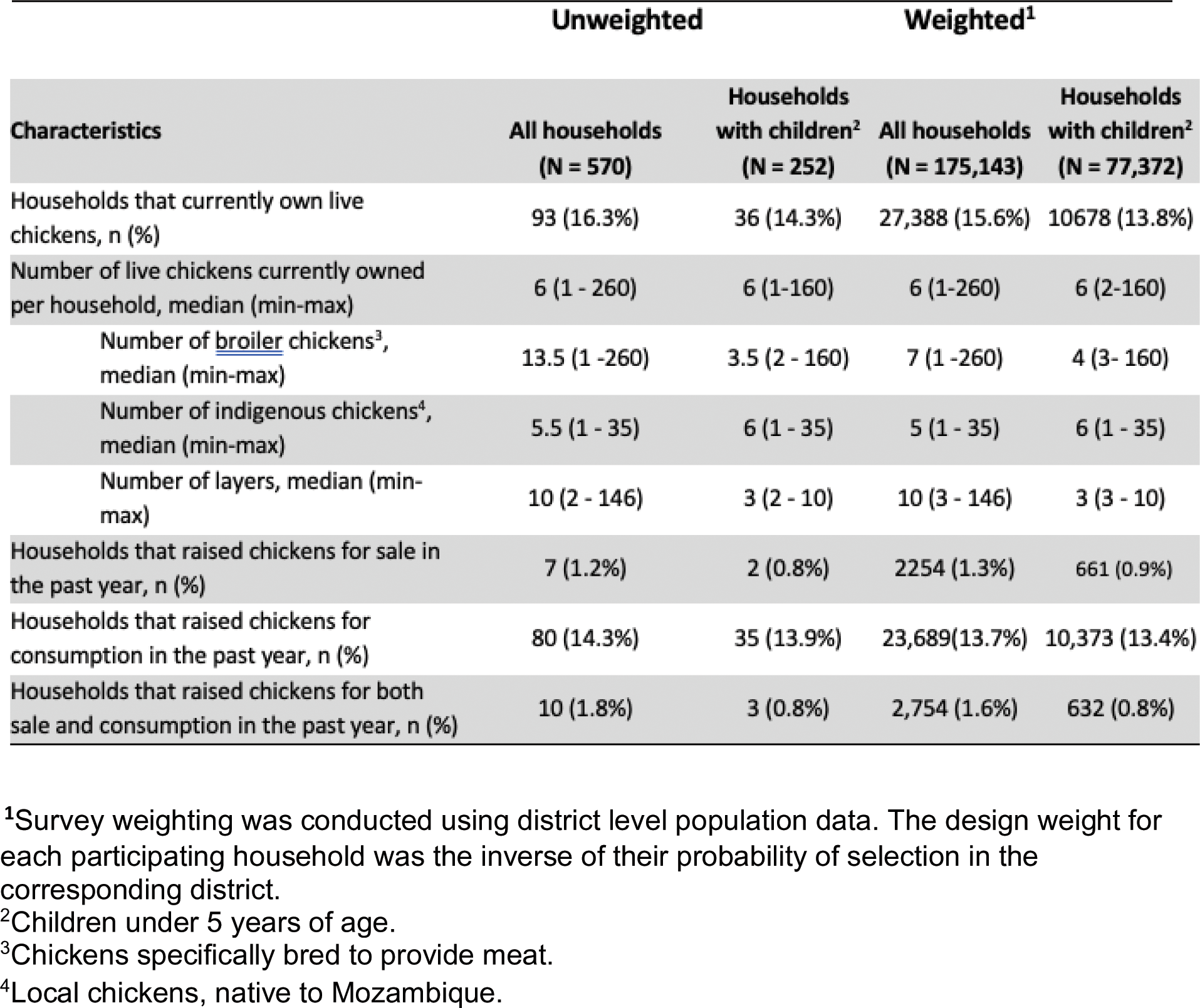
Ownership of live chickens in Maputo, Mozambique in May-June 2021.

Enumerators observed chicken feces on the floor or ground at the household or compound at 52.6% of the households that raised live chickens and 75.2% households with children under five that raised live chickens (Table 5). A small proportion of households (1.3%) reported that they kept chickens inside of their home. Storing uncooked poultry meat without refrigeration was very uncommon. Of households with children under five that raised chickens at the time of survey, 33.1% reported that their children take care of live chickens and 14.9% reported that their children collect eggs. About 20% of the households with children under five that raised live chickens provided measures to separate children from chickens.

**Table 5.**
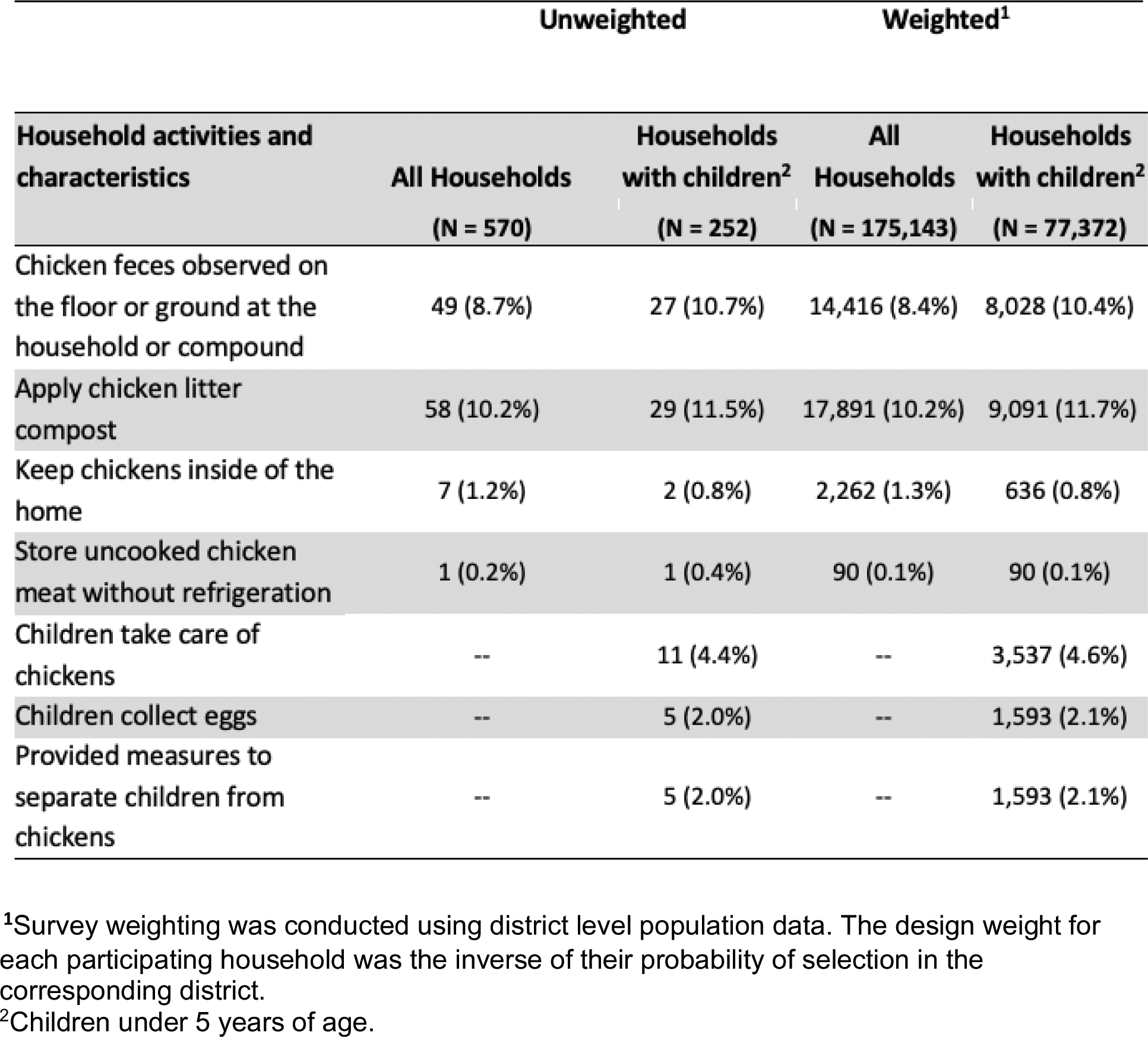
Behaviors and environment of households raising live chickens that may increase exposure to chicken feces and enteropathogens in Maputo, Mozambique in May-June 2021.

### Chicken litter compost (weighted analysis)

Applying chicken litter compost to gardens occurred but was not common (10.2%). The most common source of chicken litter compost was farmers (38.6%), followed by broiler chickens owned by households (23.7%) and family, friends, or neighbors (23.6%) (Table 3, Supplementary Table 2-4).

## DISCUSSION

Estimates of domestic purchase, ownership, and consumption of live chickens and poultry meat and eggs are sparse; here we provide such data among households in Maputo, Mozambique. Our key findings were: 1) the majority of households consumed either poultry meat or eggs or both in the previous week of the survey; and 2) the most common source of poultry was corner stores, followed by wet markets, both of which often do not have access to adequate hygiene facilities.^15^ These findings show the importance and potentially large impact of implementing food safety measures throughout the food system to control foodborne illness from chickens, which we have previously shown to be a primary driver of transmission of *Campylobacter jejuni/coli* and *Salmonella* spp..^15,33,34^ Our data on the consumption and purchasing patterns of poultry products suggest that poultry farming provides an important source of animal protein in this setting, and offer important information for identifying points of control in both Mozambique and LMICs at large. Our previous study identified a high prevalence of *Campylobacter* and *Salmonella* contamination in chicken feces and carcasses at corner stores and wet markets in Maputo.^15^ Data from this study have also been used to interpret results of a simulation model of pathogens of animal origin.^24^ The added information in this study can be used to more accurately quantify the impact of chicken-sourced pathogens on health risks and to inform the development of effective control mechanisms throughout the poultry value chain, contributing to improvements in food safety in the broader food system. The median rate of foodborne DALYs was estimated to be 1,179 per 100,000 population in 2010 among high-mortality countries in the African region, which includes Mozambique, indicating the need for better control of foodborne illness.^2^ As chickens provide a critical source of nutrition and income, and with the anticipated growth of poultry farming in LMICs in the coming decades, it is important to ensure food safety, especially among the vulnerable population.^17,25,35^

Our survey identified corner stores as the primary source of chicken products in Maputo, illustrating the potential for exposure to enteropathogens by consumers. Our previous study found that 25% of broiler chicken meat at these stores was contaminated with *Campylobacter jejuni/coli,* and 15% were contaminated with *Salmonella* spp.^15^ At wet markets, which was identified as the second most popular source of chicken products among households in Maputo, the prevalence of contamination was even higher. *C. jejuni/coli* was found in 100% of broiler chicken meat samples, and *Salmonella* spp. was identified in 17% at wet markets. These findings across our field studies underscore the urgent need for improved food safety measures at these locations to mitigate the risk of foodborne transmission of zoonotic enteropathogens. Using transmission dynamic modeling, we previously estimated that controlling foodborne transmission would significantly reduce the infection risk of these pathogens in this setting.^33^ Wet markets are also linked to the emergence of zoonotic pathogens with high potential for causing epidemics and pandemics,^34^ making it more important to discuss regulations at these locations to mitigate health risks among local communities and beyond.

The ownership of live chickens represents another critical source of exposure to enteropathogens of poultry origin. Our observations suggest that children in homes that raise chickens are commonly exposed to chicken feces, which could transmit enteropathogens in the household setting. One-fifth of the households reported having measures to separate children from chickens at home, while other households reported that children participate in taking care of chickens and collecting eggs. These observations highlight the need for household-level control measures to mitigate health risks to children while enabling them to benefit from owning live chickens at home.^37^

This study has limitations. Our survey was conducted from May to June 2021, while COVID-19 case counts were low, following a large peak from January to March 2021 in Mozambique. The global shipping crisis during the pandemic affected supplies of poultry feed, influencing poultry production in Mozambique and everywhere.^38^ Market hours were also shortened in response to the pandemic. Although we observed a decrease in chicken sales at markets, the production reached the pre-pandemic level by the time when the survey was conducted. Most questions asked about behaviors in the previous week of the survey, while a few questions asked about the past year (i.e., chicken ownership and sales), which are subject to recall bias.

Our population-based survey yielded important data on the ownership of live chickens and the purchase and consumption of poultry products among households in Maputo, Mozambique. These findings can serve as a foundation for identifying control measures at both the household and community levels. To ensure safe access to poultry products, which are the primary source of animal protein, nutrition, and income for individuals and children, enhancing food hygiene and biosafety resources at corner stores and wet markets is essential. Interventions should be designed based on insights from various disciplines (food safety, consumer science, sociology, policy, and education)^39^ with extensive input from local stakeholders. Improved regulations and infrastructure in these settings can contribute to the WHO Global Strategy for Food Safety 2022-2030^40^ and play a key role in mitigating the risk of the emergence of new pathogens with a high potential for causing epidemics and pandemics.

## Data Availability

Data produced in the present study are available upon reasonable request to the authors

## ACKNOWLEDGEMENTS

This research was supported by the Bill & Melinda Gates Foundation (OPP1189339). FL was supported by the National Institute of Allergy and Infectious Diseases of the National Institutes of Health under Award Number T32AI138952. The content is solely the responsibility of the authors and does not necessarily represent the official views of the National Institutes of Health. We thank Dr. Ian Hennessee for sharing his GIS mapping and R code.

## AUTHOR CONTRIBUTION (following the CRediT author statement requirements)

**Kayoko Shioda:** Conceptualization, Methodology, Validation, Formal Analysis, Data Curation, Writing - Original Draft, Writing - Review & Editing, Visualization; **Hermógenes Neves Mucache:** Conceptualization, Methodology, Investigation, Resources, Data Curation, Writing - Review & Editing, Supervision, Project Administration; **Frederica Lamar:** Conceptualization, Methodology, Investigation, Data Curation, Writing - Review & Editing; **Jhanel Chew:** Formal Analysis, Writing - Original Draft, Visualization, Writing - Review & Editing; **Anushka Reddy Marri:** Formal Analysis, Visualization, Writing - Review & Editing; **Karen Levy:** Conceptualization, Methodology, Resources, Writing - Review & Editing, Supervision, Project Administration, Funding Acquisition; **Matthew Freeman:** Conceptualization, Methodology, Resources, Writing - Review & Editing, Supervision, Project Administration, Funding Acquisition.

## SUPPLEMENTARY METHODS

### Modules and questions in the survey

We first gave a consent to at least one adult household member to participate in the survey. Next, we conducted the COVID-19 screening and postponed a household visit if a household failed to pass the screening questions.

There were seven modules in the survey, followed by an observational component. Information was collected for each type of poultry products (meat and egg) as well as the following type of chickens: broiler chickens (i.e., chickens specifically bred to provide meat), layer chickens, and indigenous chickens (i.e., local chickens, native to Mozambiquie). For households with children under five years of age, we specifically asked about the corresponding information for children.

In the first module (Module A), we asked about household demographics, such as the number of household members and the number of children under five.

The second module (Module B) was about the purchase and consumption of chicken meat and eggs. We asked whether households purchased chicken meat and eggs in the previous week of the survey date, and if yes, asked about the location/source where they purchased them. We asked whether households consumed chicken meat (broiler, indigenous) and/or eggs at any setting, including but not limited to the home and restaurants, and if yes, how many days in the past week they ate them. For households with children under five, we asked these questions for children as well. We also asked where households store uncooked chicken meat.

The third module (Module C) was about the history of raising chickens for sale. We asked whether households previously raised chickens for sale at their house or compound, and if yes, whether they had a chicken coop and how many flocks or groups of birds they produced last year. If household reported that they used chicken litter as bedding, we asked what they usually do with the used chicken litter (e.g., throw it away, reuse it for another flock, repackage it to sell, apply it to garden).

The fourth module (Module D) was about the current ownership of live chickens (layers, broilers, and indigenous chickens). We asked whether households currently raise live chickens for sale or consumption, and if yes, how many live chickens they currently own and where they most often keep them during the day and at night. We then asked whether households purchased live chickens for their own consumption or use in the previous week from the survey date, and if yes, we asked about the location/source of these live chickens. For households with children under five, we asked whether children help take care of live chickens. We also asked what measures, if any, households take to separate chickens from children (e.g., corralling chickens).

The fifth module (Module E) was about chicken litter compost. We asked if households had a garden, and if yes, whether they apply chicken litter compost to the garden. We also asked where they purchase or get chicken litter compost and if children help apply chicken litter compost to their garden.

The sixth module (Module F) was about water, sanitation and hygiene (WASH), and the last module (Module G) was about COVID-19. We asked whether households had any negative effect from the pandemic in their households, what kind of aid they had received if any, what kind of changes they experienced during the pandemic (e.g., frequency of market visits), hygiene practice and other personal prevention measures during the pandemic, and their perspectives on COVID-19.

In the end of the household visit, enumerators conducted an observation related to hygiene. They observed handwashing facilities, products for general cleaning, and where households keep chickens.

## SUPPLEMENTARY TABLES

**Supplementary Table 1.** Multi-stage cluster sampling strategy in Maputo, Mozambique.

| District | Population | Percentage of the total population of Maputo | Number of clusters of 20 households | Number of neighborhoods | Name of sampled neighborhoods |
| --- | --- | --- | --- | --- | --- |
| KaMavota | 293,766 | 29% | 8 | 4 | 3 de Fevereiro<br>Costa do Sol<br>Mahotas<br>Hulene B |
| KaMaxakeni | 223,936 | 22% | 6 | 3 | Urbanização<br>Maxaquene A<br>Maxaquene C |
| KaMubukwana | 304,524 | 31% | 8 | 4 | Malhazine<br>Zimpeto<br>Bagamoyo<br>Inhagoia B |
| KaTembe | 20,629 | 2% | 1 | 1 | Chali |
| Nlhamankulu | 155,462 | 16% | 4 | 2 | Chamanculo D<br>Aeroporto A |
| **Total (City of Maputo)** | **998,317** | **100%** | **27** | **14** |  |

**Supplementary Table 2.** Source of live chickens, chicken meat, eggs, and chicken litter compost among households that purchased these products in the past week of the survey date in Maputo, Mozambique in May-June 2021 (unweighted statistics).

| Source | Broiler chicken meat<br>(N=250) | Live broiler chickens<br>(N=49) | Live indigenous chicken<br>(N= 6) | Eggs<br>(N=263) | Chicken litter compost<br>(N=58) |
| --- | --- | --- | --- | --- | --- |
| Markets | 43 (17.2%) | 9 (18.4%) | 1 (16.7%) | 43 (16.3%) | 1 (1.7%) |
| Supermarkets | 7 (2.8%) | 0 (0%) | 0 (0%) | 10 (3.8%) | 0 (%) |
| Corner stores | 141 (56.4%) | 0 (0%) | 2 (33.3%) | 162 (61.6%) | 0 (%) |
| Family/friends/neighbors | 4 (1.6%) | 5 (10.2%) | 3 (50%) | 13 (5.0%) | 13 (22.4%) |
| Directly from farmers | 45 (18.0%) | 32 (65.3%) | 0 (0%) | 22 (8.4%) | 23 (39.7%) |
| Directly from layers that I own | 1 (0.4%) | 1 (2.0%) | 0 (0%) | 6 (2.3%) | 2 (3.4%) |
| Directly from broiler chickens that I own | 8 (3.2%) | 2 (4.1%) | 0 (0%) | 1 (0.4%) | 14 (24.1%) |

**Supplementary Table 3.**
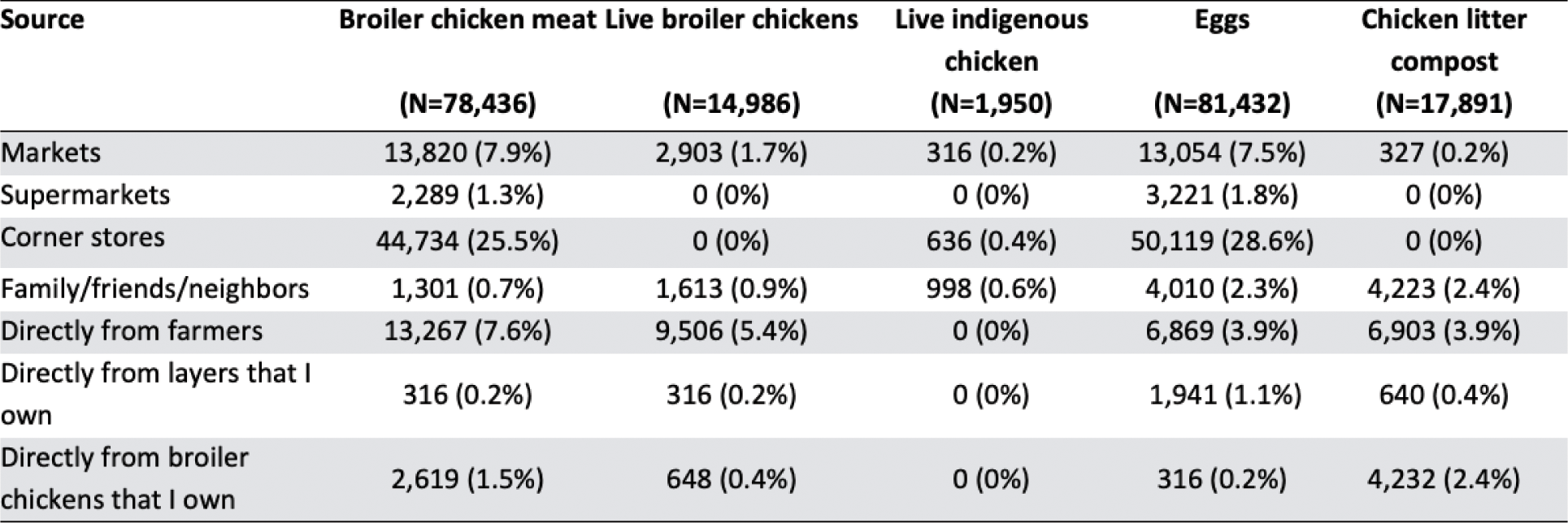
Source of live chickens, chicken meat, eggs, and chicken litter compost among households that purchased these products in the past week of the survey date in Maputo, Mozambique in May-June 2021 (weighted statistics^1^ with percentages for total households (N=175,143)).

**Supplementary Table 4.** Source of live chickens, chicken meat, eggs, and chicken litter compost among households that purchased these products in the past week of the survey date in Maputo, Mozambique in May-June 2021 (unweighted statistics with percentages for total households (N=570)).

| Source | Broiler chicken meat<br>(N=250) | Live broiler chickens<br>(N=49) | Live indigenous<br>chicken<br>(N= 6) | Eggs<br>(N=263) | Chicken litter<br>compost<br>(N=58) |
| --- | --- | --- | --- | --- | --- |
| Markets | 43 (7.5%) | 9 (1.6%) | 1 (0.2%) | 43 (7.5%) | 1 (0.2%) |
| Supermarkets | 7 (1.2%) | 0 (0%) | 0 (0%) | 10 (1.8%) | 0 (%) |
| Corner stores | 141 (24.7%) | 0 (0%) | 2 (0.4%) | 162 (28.4%) | 0 (%) |
| Family/friends/neighbors | 4 (0.7%) | 5 (0.9%) | 3 (0.5%) | 13 (2.3%) | 13 (2.3%) |
| Directly from farmers | 45 (7.9%) | 32 (5.6%) | 0 (0%) | 22 (3.9%) | 23 (4.0%) |
| Directly from layers that I own | 1 (0.2%) | 1 (0.2%) | 0 (0%) | 6 (1.1%) | 2 (0.4%) |
| Directly from broiler chickens that I own | 8 (1.4%) | 2 (0.4%) | 0 (0%) | 1 (0.2%) | 14 (2.5%) |

## REFERENCES

1 FAO AP and HD (AGA). Shaping the future of livestock: Sustainably, responsibly, efficiently. Rome: FAO, 2018 http://www.fao.org/publications/card/en/c/I8384EN/ (accessed April 22, 2021).

2 Havelaar AH, Kirk MD, Torgerson PR, et al. World Health Organization Global Estimates and Regional Comparisons of the Burden of Foodborne Disease in 2010. PLOS Med 2015; 12: e1001923.

3 Murray CJ, Vos T, Lozano R, et al. Disability-adjusted life years (DALYs) for 291 diseases and injuries in 21 regions, 1990–2010: a systematic analysis for the Global Burden of Disease Study 2010. Lancet 2013; 380: 2197–223.

4 Gay-Andrieu E, Adehossi E, Illa H, Garba Ben A, Kourna H, Boureima H. [Prevalence of cryptosporidiosis in pediatric hospital patients in Niamey, Niger]. Bull Soc Pathol Exot 1990 2007; 100: 193–6.

5 Checkley W, Gilman RH, Epstein LD, et al. Asymptomatic and symptomatic cryptosporidiosis: their acute effect on weight gain in Peruvian children. Am J Epidemiol 1997; 145: 156–63.

6 Platts-Mills JA, Liu J, Rogawski ET, et al. Use of quantitative molecular diagnostic methods to assess the aetiology, burden, and clinical characteristics of diarrhoea in children in low-resource settings: a reanalysis of the MAL-ED cohort study. Lancet Glob Health 2018; 6: e1309–18.

7 Ngure FM, Humphrey JH, Mbuya MNN, et al. Formative Research on Hygiene Behaviors and Geophagy among Infants and Young Children and Implications of Exposure to Fecal Bacteria. Am J Trop Med Hyg 2013; 89: 709–16.

8 Ngure F, Gelli A, Becquey E, et al. Exposure to Livestock Feces and Water Quality, Sanitation, and Hygiene (WASH) Conditions among Caregivers and Young Children: Formative Research in Rural Burkina Faso. Am J Trop Med Hyg 2019; 100: 998–1004.

9 Reid B, Orgle J, Roy K, Pongolani C, Chileshe M, Stoltzfus R. Characterizing Potential Risks of Fecal-Oral Microbial Transmission for Infants and Young Children in Rural Zambia. Am J Trop Med Hyg 2018; 98: 816–23.

10 Ercumen A, Prottas C, Harris A, Dioguardi A, Dowd G, Guiteras R. Poultry Ownership Associated with Increased Risk of Child Diarrhea: Cross-Sectional Evidence from Uganda. Am J Trop Med Hyg 2020; 102: 526–33.

11 Zambrano LD, Levy K, Menezes NP, Freeman MC. Human diarrhea infections associated with domestic animal husbandry: a systematic review and meta-analysis. Trans R Soc Trop Med Hyg 2014; 108: 313–25.

12 Jones AD, Colecraft EK, Awuah RB, et al. Livestock ownership is associated with higher odds of anaemia among preschool-aged children, but not women of reproductive age in Ghana. Matern Child Nutr 2018; 14: e12604.

13 Headey D, Nguyen P, Kim S, Rawat R, Ruel M, Menon P. Is Exposure to Animal Feces Harmful to Child Nutrition and Health Outcomes? A Multicountry Observational Analysis. Am J Trop Med Hyg 2017; 96: 961–9.

14 WHO | WHO estimates of the global burden of foodborne diseases. WHO. http://www.who.int/foodsafety/publications/foodborne_disease/fergreport/en/ (accessed Sept 16, 2020).

15 Lamar F, Mucache HN, Mondlane-Milisse A, et al. Quantifying Enteropathogen Contamination along Chicken Value Chains in Maputo, Mozambique: A Multidisciplinary and Mixed-Methods Approach to Identifying High Exposure Settings. Environ Health Perspect 2023; 131: 117007.

16 Berendes DM, Yang PJ, Lai A, Hu D, Brown J. Estimation of global recoverable human and animal faecal biomass. Nat Sustain 2018; 1: 679–85.

17 Wong JT, de Bruyn J, Bagnol B, et al. Small-scale poultry and food security in resource-poor settings: A review. Glob Food Secur 2017; 15: 43–52.

18 UN General Assembly. Transforming our world : the 2030 Agenda for Sustainable Development. 2015.

19 Melesse A. Significance of scavenging chicken production in the rural community of Africa for enhanced food security. Worlds Poult Sci J 2014; 70: 593–606.

20 Bank AD. Resumo dos Resultados 2018 – Moçambique. Afr. Dev. Bank - Build. Today Better Afr. Tomorrow. 2019; published online May 10. https://www.afdb.org/en/documents/document/resumo-dos-resultados-2018-mocambique-106488 (accessed Jan 23, 2023).

21 Ministério da Agricultura e Segurança Alimentar. Anuário de Estatísticas Agrárias 2015. https://www.masa.gov.mz/wp-content/uploads/2017/12/Anuario_Estatistico2016.pdf (accessed Jan 23, 2023).

22 Vipham JL, Amenu K, Alonso S, et al. No food security without food safety: Lessons from livestock related research. Glob Food Secur 2020; 26: 100382.

23 Kaur M, Graham JP, Eisenberg JNS. Livestock Ownership Among Rural Households and Child Morbidity and Mortality: An Analysis of Demographic Health Survey Data from 30 Sub-Saharan African Countries (2005-2015). Am J Trop Med Hyg 2017; 96: 741–8.

24 Bardosh KL, Hussein JW, Sadik EA, et al. Chicken eggs, childhood stunting and environmental hygiene: an ethnographic study from the Campylobacter genomics and environmental enteric dysfunction (CAGED) project in Ethiopia. One Health Outlook 2020; 2: 5.

25 Alders RG, Dumas SE, Rukambile E, et al. Family poultry: Multiple roles, systems, challenges, and options for sustainable contributions to household nutrition security through a planetary health lens. Matern Child Nutr 2018; 14: e12668.

26 Scudiero L, Tak M, Alarcón P, Shankar B. Understanding household and food system determinants of chicken and egg consumption in India. Food Secur 2023; 15: 1231–54.

27 Comercialização de Carnes. Américo Da Conceição PDF | PDF | Matadouro | Avicultura. Scribd. https://pt.scribd.com/document/472833953/Comercializacao-de-carnes-Americo-da-Conceicao-pdf (accessed March 3, 2024).

28 Bah E, Gajigo O. Improving the Poultry Value Chain in Mozambique. https://www.afdb.org/fileadmin/uploads/afdb/Documents/Publications/WPS_No_309_Improving_the_Poultry_Value_Chain_in_Mozambique.pdf (accessed Jan 23, 2023).

29 Maputo Cidade — Instituto Nacional de Estatistica. http://www.ine.gov.mz/iv-rgph-2017/maputo-cidade (accessed Jan 23, 2023).

30 McKague K, Karnani A. Job Creation in the Mozambican Poultry Industry. Financ Rev 2014; published online Jan 1.

31 Lin B, Dietrich ML, Senior RA, Wilcove DS. A better classification of wet markets is key to safeguarding human health and biodiversity. Lancet Planet Health 2021; 5: e386–94.

32 Lumley T. survey: analysis of complex survey samples. 2023.

33 Shioda K, Brouwer AF, Lamar F, Mucache HN, Levy K, Freeman MC. Opportunities to Interrupt Transmission of Enteropathogens of Poultry Origin in Maputo, Mozambique: A Transmission Model Analysis. Environ Health Perspect; 131: 117004.

34 Naguib MM, Li R, Ling J, Grace D, Nguyen-Viet H, Lindahl JF. Live and Wet Markets: Food Access versus the Risk of Disease Emergence. Trends Microbiol 2021; 29: 573–81.

35 Chaiban C, Robinson TP, Fèvre EM, et al. Early intensification of backyard poultry systems in the tropics: a case study. Animal; 14: 2387–96.

36 Bardosh KL, Hussein JW, Sadik EA, et al. Chicken eggs, childhood stunting and environmental hygiene: an ethnographic study from the Campylobacter genomics and environmental enteric dysfunction (CAGED) project in Ethiopia. One Health Outlook 2020; 2. DOI:10.1186/s42522-020-00012-9.

37 Passarelli S, Ambikapathi R, Gunaratna NS, et al. The role of chicken management practices in children’s exposure to environmental contamination: a mixed-methods analysis. BMC Public Health 2021; 21: 1097.

38 Palouj M, Lavaei Adaryani R, Alambeigi A, Movarej M, Safi Sis Y. Surveying the impact of the coronavirus (COVID-19) on the poultry supply chain: A mixed methods study. Food Control 2021; 126: 108084.

39 Langsrud S, Veflen N, Allison R, et al. A trans disciplinary and multi actor approach to develop high impact food safety messages to consumers: Time for a revision of the WHO - Five keys to safer food? Trends Food Sci Technol 2023; 133: 87–98.

40 WHO global strategy for food safety 2022-2030: towards stronger food safety systems and global cooperation. https://www.who.int/publications-detail-redirect/9789240057685 (accessed March 1, 2024).

